# Improving cardiovascular health in patients with an abdominal aortic aneurysm: Development of the Cardiovascular risk reduction in patients with aneurysms (CRISP) behaviour change intervention

**DOI:** 10.1101/2022.12.14.22283464

**Authors:** T. M. Withers, C. J. Greaves, M. J. Bown, A. Saratzis

## Abstract

**Objectives:** Abdominal aortic aneurysm (AAA) is an important cardiovascular health problem. Ultrasound screening is proven to reduce AAA-mortality and programmes have been implemented in some healthcare systems. Those who are identified as having a small AAA in screening enter into a surveillance programme to monitor AAA-size. Individuals in AAA-surveillance are at elevated risk of cardiovascular events, which is not currently addressed sufficiently. We aimed to develop a simple intervention to reduce cardiovascular risk, which could be embedded in AAA surveillance-pathways.

**Design:** Intervention mapping methods were used to co-develop the intervention

**Participants:** Individuals with AAA, families/carers, and healthcare staff.

**Setting:** AAA screening and surveillance programmes in the United Kingdom

**Outcome measures:** We identified “targets for change” by synthesising research evidence and international guidelines and consulting with patients, caregivers and health service providers. We conducted a series of workshops to identify barriers to and facilitators of change, and used taxonomies of behaviour change theories and techniques to match intervention strategies to each target. Further stakeholder involvement work helped refine the intervention.

**Results:** The developed intervention focusses on assessment and individually tailored discussion of risk factors, exchanging information, building motivation and action planning, followed by review of progress and problem-solving. Workbooks covering physical activity, diet, stress management, alcohol, smoking, blood pressure and mental health are provided to support behaviour change. The intervention is facilitated by trained healthcare professionals during the patient’s AAA screening appointment for the duration that they are in surveillance.

**Conclusions:** The developed intervention will now be tested to assess whether it can be integrated with the current AAA screening programme. The developed intervention is a novel approach to reducing cardiovascular disease in the AAA population, it is also the first intervention which tries to do this in this population.

## INTRODUCTION

All men in the UK are invited for an ultrasound scan to screen for Abdominal Aortic Aneurysm (AAA) in the year of their 65th birthday; similar programmes exist in other countries(1-4). The vast majority of those diagnosed with AAA through screening do not require immediate surgery to treat the AAA(1, 2, 4-9). They are entered into a disease-specific surveillance programme to monitor AAA growth with repeat ultrasound measurements(8, 9). Whilst screening reduces AAA-related mortality by providing an opportunity for timely surgical intervention, it has very minimal effect on all-cause mortality(2, 4, 6, 10).

Patients with AAA have an elevated cardiovascular risk; in fact, cardiovascular events are one of the principle causes of morbidity and mortality amongst those in AAA surveillance (2, 4, 6, 7). This elevated risk is mainly driven by modifiable risk factors such as smoking and excess weight (2, 4, 6).

The regular attendance of these individuals with AAA at surveillance clinics represents an excellent opportunity to address their excess cardiovascular-risk within an existing, well defined, and well attended clinical pathway. AAA-surveillance pathways were not specifically developed to deliver cardiovascular-risk modification. Further those with AAA have unique characteristics that make the adoption of cardiovascular-interventions developed in different clinical settings challenging. They typically suffer from multiple co-morbidities, avoid contact with primary or secondary healthcare (even though AAA-surveillance attendance consistently exceeds 80%), have poor medication adherence, and are often socio-economically deprived (2-4, 11, 12). Consequently, uptake of cardiovascular-risk management has been virtually non-existent in AAA surveillance, despite some attempts by screening programmes to offer cardiovascular-risk reduction interventions (2-4, 11, 13). No high-quality research has been undertaken to develop and test interventions to reduce cardiovascular-risk in this clinical area.

Given the excellent attendance and low dropout of AAA-surveillance, this is a major opportunity to offer better cardiovascular prevention to a population at very high risk for cardiovascular events (8, 9). This research addressed this opportunity by developing a cardiovascular risk reduction intervention specifically designed to be embedded within the existing NHS Abdominal Aortic Aneurysm Screening Programme (NAAASP) clinical care pathway, hoping to reduce the chance of cardiovascular events following the diagnosis of an AAA.

## METHODS AND RESULTS

This research was approved by the NHS Wales Research Ethics Committee 7 (NHS Wales REC 7) and the NHS Health Research Authority (HRA) in January 2020 (reference: 19/EM/0366). The research was funded by the National Institute for Health and Care Research (NIHR) Academy (reference: NIHR300059) and sponsored by the University of Leicester (reference: 0479); the funder and sponsor had no input in data collection, analysis or interpretation. Participants provided written informed consent upon recruitment.

The main aim was to develop a complex clinical intervention, the CRISP intervention, in order to reduce cardiovascular risk in those taking part in the NHS AAA surveillance programmes across the UK and who have a small or medium size aneurysm. Based on current definitions, a small or medium AAA measures between 3.0 and 5.5cm in maximal anteroposterior diameter (ultrasound measurement), using NAAASP standard operating procedures (SOPs) (2, 5). Patients with an AAA exceeding 5.5cm in size are referred to secondary care for consideration of surgical repair(1, 8, 9, 14, 15). All patients are advised to take an antiplatelet and lipid-control medications (statin); however the mechanism to ensure this happens varies considerably across screening programmes (1, 2, 6, 8).

The CRISP intervention was designed based on the Medical Research Council (MRC) guidelines for the development of complex interventions for clinical settings (16), using the intervention mapping approach and principles. The intervention mapping framework, widely used in the development of health behaviour change interventions, is described in detail elsewhere (17). It comprises a six-step ecological approach to assessing and intervening in health problems via the development or modification of an intervention.

- Step 1: Logic model of the problem
- Step 2: Program outcomes and objectives – Logic model of change
- Step 3: Program design
- Step 4: Program production
- Step 5: Program implementation plan
- Step 6: Evaluation plan

As this is an iterative process, methods and results will be presented together for each step.

### Patient and public involvement

Patients were involved throughout the development of the intervention and formed one of the studies advisory groups.

### Participants and stakeholders

To develop the intervention, three stakeholder groups were formed:

Group A: Patient (lay) advisory group, consisting of 63 men with a small or medium sized AAA who were taking part in AAA surveillance within NAAASP, 26 men who had undergone AAA repair surgery following attendance to AAA screening in the NHS (AAA picked up via AAA screening), 8 men who had been considered for AAA repair once they had been diagnosed with an AAA via screening and had attended at least one surveillance appointment but were eventually deemed not fit for surgery once they had been seen in secondary care, one man who had surgery due to an AAA rupture and had survived (had previously not attended screening despite having received an invitation), and one man diagnosed with an AAA after having suffered a myocardial infarction (incidental finding) and awaiting to be seen in AAA surveillance via NAAASP. Seven partners (wives in all cases) also took part.

Group B: A service provider advisory group, consisting of 43 healthcare staff involved in delivery and management of NHS AAA screening/surveillance.

Group C: A cardiovascular expert advisory group, consisting of 21 clinicians involved in cardiovascular care of individuals with AAA (cardiologists, vascular surgeons, general practitioners, pharmacists, vascular nurses) alongside 4 experts on the prescription of exercise to reduce cardiovascular risk.

Further to the 3 research-specific advisory groups, advice was sought, when necessary from the NAAAP Research Committee, NAAASP clinical lead, a senior Public Health (Professor Holland, University of Leicester) and Behaviour Change (Professor Colin Greaves) researcher, as well as members of the councils of the Vascular Society of Great Britain and Ireland (VSGBI) and Society of Vascular Nursing. An independent AAA-specific patient advisory group provided advice on patient facing materials, barrier, and facilitators. Participants were recruited to the stakeholder advisory groups by advertising the opportunity on social media, word of mouth and the authors asking their professional networks to circulate an advert (direct recruitment). Due to the Coronavirus Disease 2019 (COVID19) pandemic, all advisory groups were held online using videoconferencing software. The chief investigator (AS) liaised with participants who did not have access to videoconferencing to provide appropriate software and hardware (free) and facilitate participation. No lay participants or expert stakeholders invited to take part declined taking part due to inability to join the online advisory groups. Potential participants who could not understand written and spoken English were not able to take part as there was no translation facility during the advisory groups and discussion in English was necessary in order to obtain relevant data of sufficient quality. When necessary, lay individuals or experts were interviewed by AS and/or TW over the telephone to address queries.

#### Step 1: Logic Model of the Problem

To identify target behaviours for reduction cardiovascular risk the existing evidence-base on lifestyle intervention components and behaviour change strategies associated with effectiveness for reducing cardiovascular-risk was reviewed (AS, TW, CG). This included updating a prior review on this topic (18) as part of a commissioned “state of the art” review(19). We also reviewed cardiovascular-interventions developed for other high-risk populations with similar characteristics, such as male long-distance truck drivers and patients with psoriasis(20-22) and undertook a systematic review on long term effectiveness of physical activity interventions, searched other relevant systematic reviews (18) and relevant clinical guidelines (23-28). Each strategy identified was discussed with all PPI and expert stakeholders, including the likely feasibility /acceptability of using digital health interventions (e.g. mobile phone apps). Almost all lay participants were against the exclusive use of digital technologies to deliver the intervention, although they were supportive of having links to sources of online support for those who were capable and willing to use them.

#### Scaffolding questionnaire

Based on the initial work, described above, we developed a ‘scaffolding questionnaire.’ This was used to inform the initial design of the intervention. There were two versions: a patient and healthcare professional/expert version. The questions explored optimal contents as well as barriers and enablers in creating and delivering the intervention. They were modified appropriately in each group to ensure that they were relevant for the target audience. The questions focused on understanding the respondent’s knowledge of AAA, to ensure that the intervention is informative and not repeating what is already known, and their views on what cardiovascular risk factors are the most important to focus on and which one are more likely to be successful, an example scaffolding questionnaire is provided in appendix 1

A total of 14 responses were received for the patient questionnaire, made up of 10 respondents who are currently undergoing AAA screening, 3 who had and AAA repaired surgically and one who did not specify. We received 59 responses for the healthcare professional questionnaire with the most common profession of responses being from General Practitioners (GPs) and Consultant Vascular Surgeons (both making up 22% of overall responses). Both groups, patient and healthcare professionals, broadly agreed on including: medication review, increasing physical activity and heart healthy eating/weight management (**Error! Reference source not found**.). The other potential components received a more mixed response.

The scaffolding questionnaire also explored the optimum delivery method (**Error! Reference source not found**.); this information was used in step 2, described later.

Following this process we identified the below components would make up the intervention and that patients would be able to choose which components they wished to focus on:

- Stop smoking support
- Increasing physical activity
- Improving diet/weight management
- Medication review
- Managing stress/anxiety
- Managing low mood/depression
- Alcohol support, this was added retrospectively during step 2. As during the advisory groups it became apparent that a number of participants felt that this was important.

### Step 2: Programme outcomes and objectives

We then convened three online advisory group discussions with our lay membership, one joint between our cardiovascular and delivery experts, one joint between the delivery and lay group and one for the lay group, an example topic guide is presented in appendix 2. Following that, we involved primary care doctors through a national online survey (194 responses by general practitioners and practice nurses across the UK) and interviewing four GPs. We explored the following topics:

- Which are the main barriers or facilitators in engaging with cardiovascular-risk modification during AAA-surveillance? Both from the perspective of healthcare professionals and patients.
- Which of the identified barriers are modifiable and have greatest scope for change?

Based on the specification of the behavioural, environmental and psychological targets for change which emerged from the needs assessment (step 1), performance objectives were specified. Performance objectives are the description of the specific behaviours that the at-risk group or other agents (HCPs in this case) have to perform to achieve the desired change (17). For each performance objective, modifiable determinants of change were identified using several methods:

We then ‘mapped’ the performance objectives and change techniques onto strategies to change behaviour to produce intervention maps, also known as intervention matrices. An extract of the reducing alcohol consumption intervention map is presented in Table 3 with the performance objectives and modifiable determinants filling the first two columns. In addition to this the proposed mechanism of action was also identified to aid in the identification of potential behaviour change techniques (step 3) from the Theory and Techniques Tool, developed by Johnston and colleagues (33) and a taxonomy of behaviour change techniques (34). A total eight of intervention maps were developed:

**Table 1:**
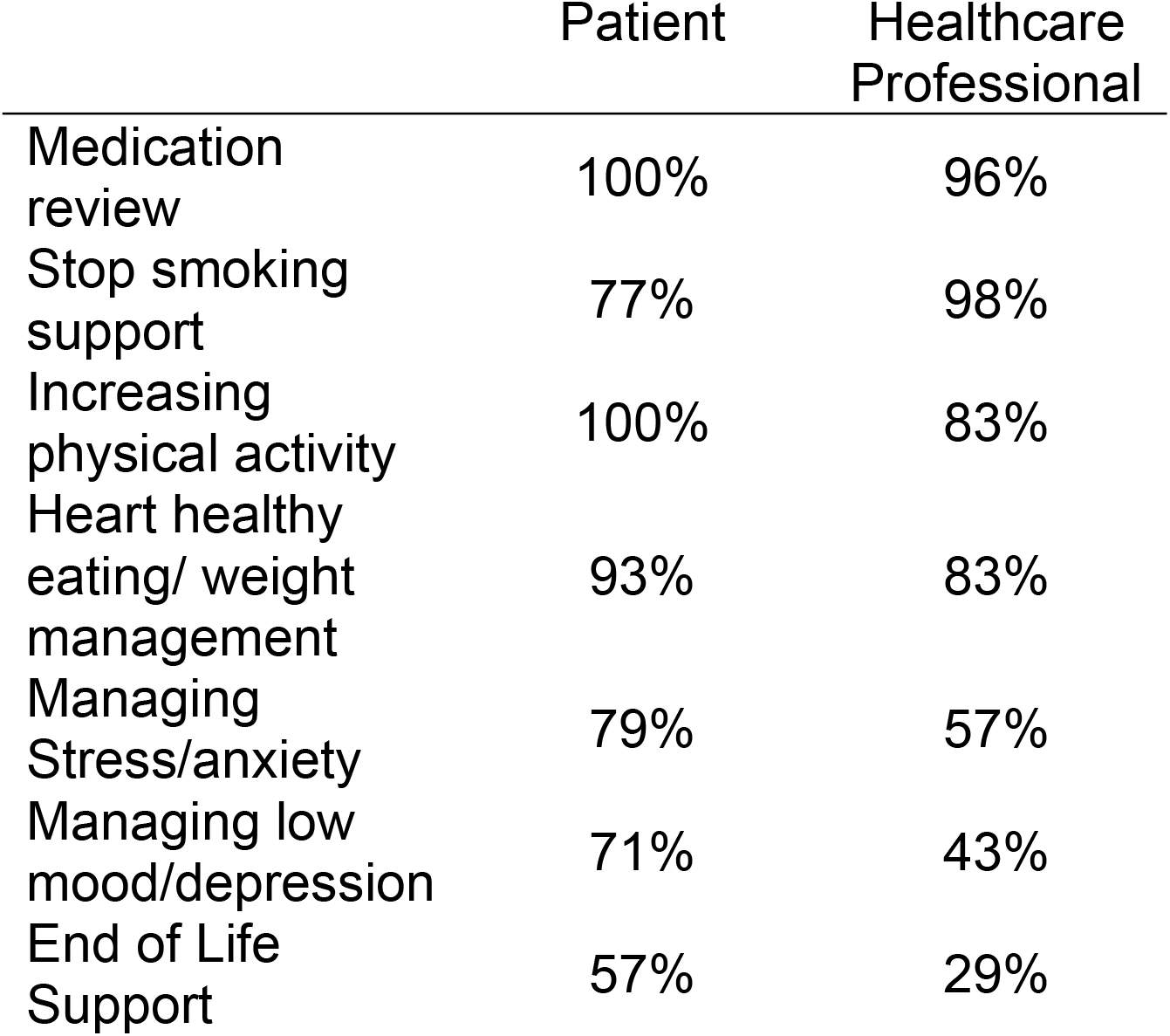
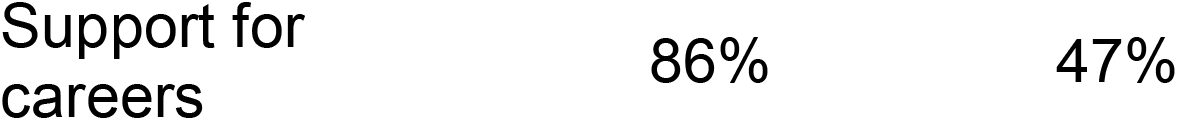
Comparison of Patient and Healthcare Professional responses. On should the component be included in the intervention. The percentage of yes responses are presented.

|  | Patient | Healthcare Professional |
| --- | --- | --- |
| Medication review | 100% | 96% |
| Stop smoking support | 77% | 98% |
| Increasing physical activity | 100% | 83% |
| Heart healthy eating/ weight management | 93% | 83% |
| Managing Stress/anxiety | 79% | 57% |
| Managing low mood/depression | 71% | 43% |
| End of Life Support | 57% | 29% |

**Table 2:** Rating the likelihood of success and implementation for different delivery methods. Where 10 represents the greatest likelihood of success.

|  | Likelihood of working |  | Ease of implementation |  |
| --- | --- | --- | --- | --- |
|  | Patient | Healthcare Professional | Patient | Healthcare Professional |
| Digital/'do it yourself' (self-care) advice and planning support | 5.0 | 3.9 | 7.6 | 6.2 |
| Paper based/self-care advice and planning support | 6.3 | 4.9 | 8.7 | 7.6 |
| 4-6 face to face meetings | 7.9 | 7.7 | 6 | 4.8 |
| 4-6 group meetings | 7 | 7.1 | 5.5 | 5.4 |
| Facilitated digital (e.g. 1 face to face meeting and 2-3 phone calls) | 7.1 | 6.2 | 7.7 | 6 |
| Facilitated paper based | 5.3 | 5.3 | 7.3 | 6.2 |
| Patients choose their preferred option (from any of the above) | 8.1 | 6.9 | 7.5 | 4.7 |

**Table 3:** Extract from the Reducing alcohol consumption intervention map

| Performance Objective | Modifiable Determinant | Change Techniques | Strategies |
| --- | --- | --- | --- |
| Accessing and engaging with support for reducing alcohol consumption.<br>This may include (a) referral to the local drug and alcohol support service if criteria are met, (b) provide self-guided support on reducing alcohol consumption via a website or booklet, (c) provide signposting to local third sector support services. | Knowledge (patient) which include:<br>1. Understanding of safe drinking levels.<br>2. Alcohol consumption is not associated with health risks.<br>Mechanism of action (MOA): Knowledge | 2.2: Feedback on behaviour<br>2.6: Biofeedback<br>5.1: Information about health consequences<br>5.3 Information about social and environmental consequences | **HCP delivered strategies:**<br>2.2: During the initial nurse assessment alcohol consumption will be assessed. Patients will be informed if their current drinking is within recommended 'healthy' levels.<br>2.6: A simple test will be offered to those who drink excessively.<br>**Information materials and digital support:**<br>5.1 and 5.3: Patients will be provided with a booklet/website with information about safe drinking levels, the social, environmental and health effects of excessive alcohol consumption, where to find help in reducing their alcohol consumption. |

1. Reducing alcohol consumption
2. Improving diet and losing weight
3. Engaging with the CRISP intervention
4. Increasing physical activity
5. Managing blood pressure
6. Smoking cessation
7. Managing stress, anxiety and low mood

In addition to the intervention maps a logic model was developed, figure 1, which shows how the intervention should work in theory.

**Figure 1:**
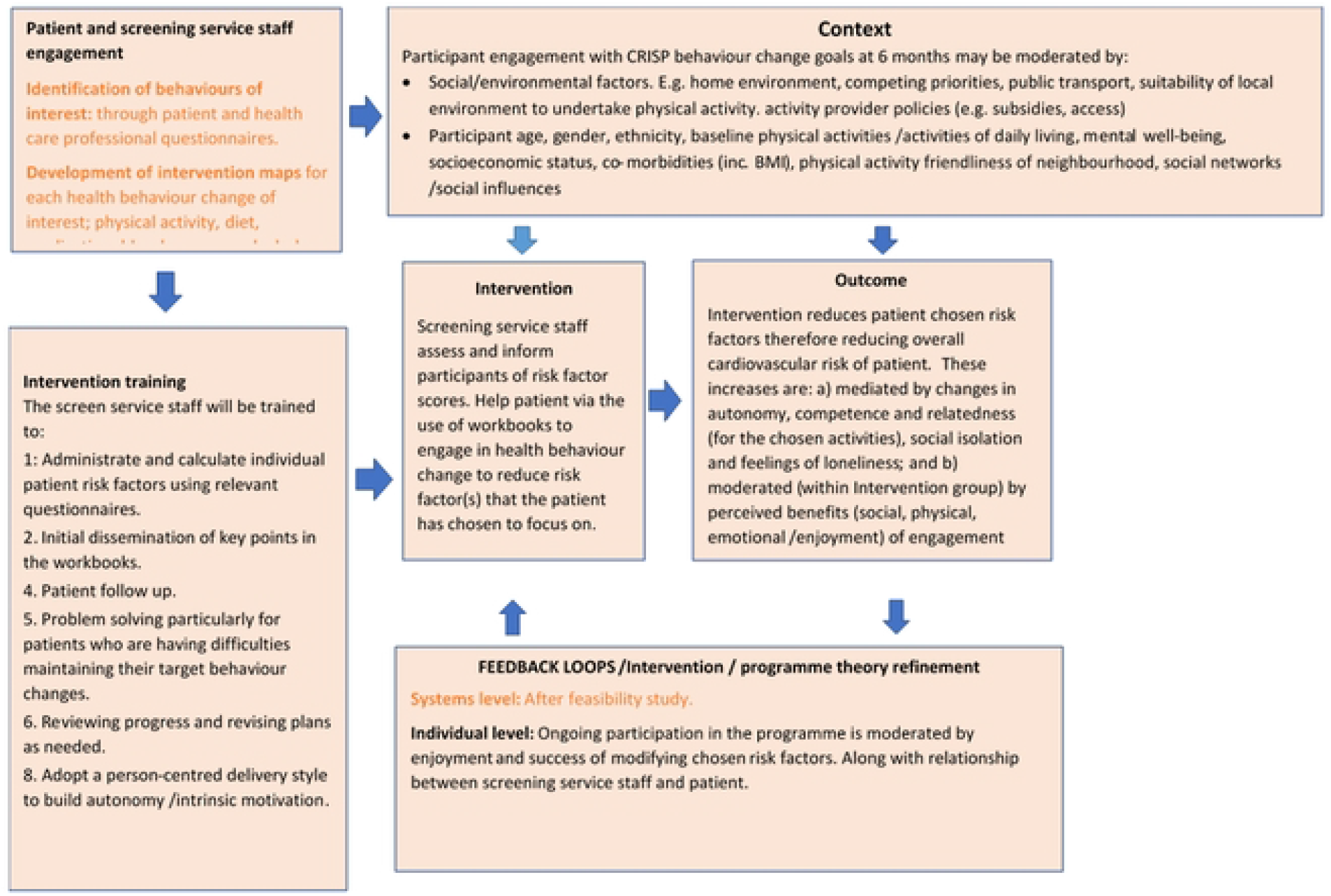
Logic model of CRISP intervention.

#### Step 3: Programme design

The third step of the intervention mapping process involved the selection of change techniques. This was done by using a combination of experience, advisory group participant suggestions and use of the behaviour change technique taxonomy (35) and theory and techniques tool (33). The latter being a way of linking behaviour change techniques with mechanisms of action. Initially all possible behaviour change techniques were identified and then reviewed by both our intervention development team (CG, TW) and by discussion with our stakeholder advisory groups. The ones which were inappropriate or not practical in the context of this study were removed. For example, ‘Instruction of how to perform behaviour’ was initially identified as potential technique to improve ‘knowledge’ for the alcohol intervention map. However, it was subsequently removed as instructions on how to drink less alcohol would have been simplistic and perceived as patronising by patients. A

#### Step 4: Programme production

The aim of this step is to finalise what the intervention is going to ‘look like’ and how the component change techniques will be organised and delivered in practice. This was initially done by recording ideas in the final column of the intervention (Table 3) about strategies that could be used to carry out the behaviour change technique. For example the strategy used for the behaviour change technique ‘feedback on behaviour’ was to give patients feedback on their drinking habit via a validated alcohol-intake questionnaire. Following this process, a draft intervention was presented to a joint meeting of all the advisory groups (lay and expert participants) to elicit their feedback and finalise the intervention format / contents.

For the final feedback session of the joint advisory group the session focused on feedback on an example work booklet, which was for physical activity. The feedback was broadly positive however a few improvements were suggested:

- Need to explain why lifting heavy weights is inappropriate but yoga and tai chi are fine.
- Offering the ability to monitor blood pressure would be beneficial as a motivational tool.
- Need to highlight what heavy household items are/are not acceptable to lift.

Following this the intervention was finalised. The finalised intervention, summarised in figure 2, is delivered in two stages. In the first stage the patient fills out a risk factor questionnaire and sends it back to their local screening service. When the screening service receive the filled-out questionnaire they enter the data into a bespoke computer programme which calculates the personalised risk factor profile of the patient this is presented as a letter.

**Figure 2:**
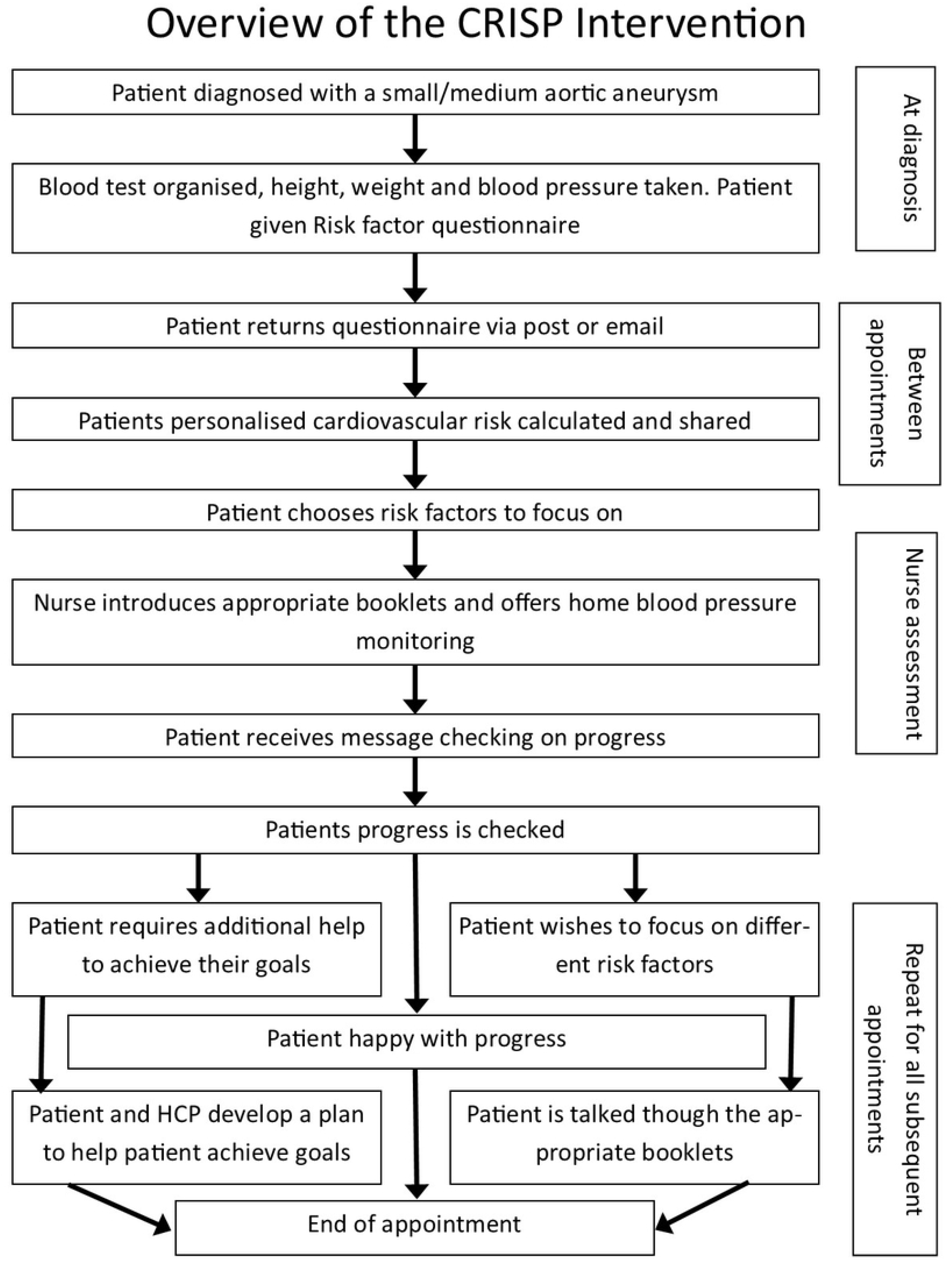
Flow chart of CRISP intervention.

The beginning of the second stage starts at the nurse assessment. The nurse assessment is part of standard care following diagnosis of a small or medium aneurysm. The purpose of the nurse assessment is to assess the overall health of the patient and suggests things a patient could do to improve their health for example; lose weight, quit smoking and or increase physical activity. The aim of the intervention is not to modify what is currently being done during the nurse assessment, which varies across the country, but to add to it.

Whilst the patient is in the waiting room waiting for the start of their nurse assessment they will be given their personalised rick factor letter to read. A small number of patients request that their nurse assessment happens via email, phone or online. In this case the letter will be sent to them beforehand. The patient will then be called in and initially their feelings and thoughts about the risk factor letter will be discussed. The patient will then be asked to pick one or two (maximum three) risk factors to focus on. The nurse may assist the patient in deciding which risk factors to focus on but it must ultimately be the patient’s decision. The nurse will then introduce the appropriate workbook(s) for the risk factors the patient has decided to focus on. A workbook has been developed for each risk factor for patients to help patients. The workbooks contains both information on why reducing the risk factor is beneficial and activities designed to help patients reduce the risk factor. In subsequent appointments, which alternate between phone and in person contacts initially patient progress will be reviewed. This will be followed by problem solving collaboratively with the patient any challenges they may have come across. This may include but is not limited to modifying goals, working through solutions to identified barriers and changing risk factors to focus on. Finally, the session will review and set goals until the next appointment. All patient contacts should last no longer than 30 minutes on average. The length of time between face to face appointments is dictated by the AAA screening programme, with the phone appointments happening approximately half way between the face to face appointments.

### Steps 5 and 6: Program Implementation and Evaluation Plan

Implementation planning is built into our approach via the involvement of service providers and service managers as stakeholders in the co-development of the intervention. The intervention has been specifically designed to be feasible for implementation within the NHS AAA Screening Programme. Feedback from the planned feasibility study and trial (below) will help to iteratively refine and optimise implementation.

Evaluation of the CRISP intervention will involve a feasibility trial including a mixed-methods process evaluation focused on refining the intervention process model (as well as establishing acceptability and fidelity of the intervention and trial procedures). The aim of the process evaluation is twofold: to test the logic model and assess intervention fidelity This is currently ongoing. Following this, the intervention will be modified based on the feedback received and a version 2 developed. Following this a multi-site randomised controlled trial to establish effectiveness and cost-effectiveness will follow and, if successful, the intervention will be rolled out across the AAA surveillance programme.

The final step of intervention mapping is to develop an in-service evaluation plan when the intervention has been adopted to ensure constant improvement. This step will be completed if/when the intervention is rolled out across NAAASP.

## DISCUSSION

In work package 1 of this research we developed a complex clinical intervention which is purpose-built for use in AAA screening and surveillance pathways, aiming to reduce the cardiovascular risk of the participants. The intervention is based not only on best available evidence /guidance and on behaviour change theory, but most importantly it is grounded on the views and opinions of patients and healthcare professionals. The final intervention, currently being feasibility-tested in the NHS, consists of a number of workbooks and interactions between NAAASP staff and patients.

The intervention mapping framework used to develop the intervention was time and resource intensive, requiring one research fellow to work on it full time for 1.5 years, but produced an intervention for use in clinical care that take into consideration the requirements of the patient, healthcare professionals and service providers. The core research team required to develop this intervention consisted of two vascular surgeons (AS, MJB), two experienced psychologists with expertise in intervention mapping (CG, TW), and a national support network of experts in a variety of clinical areas. Further, hundreds of lay individuals (with or without aneurysms) supported our advisory groups, interviews, and online surveys. The resources required to develop this intervention were considerable, with an overall cost of £244,000 for the development activities (funded by the NIHR). At the same time, there are 348 preventable cardiovascular-deaths and 720 non-fatal major cardiovascular-events every year in England amongst men in AAA surveillance. A 20% decrease would lead to an annual saving of £14.8 million in treatment costs alone across the NHS (3, 8, 9, 12, 36). Achieving a systolic blood pressure <140mmHg, stopping smoking and achieving normal LDL-levels in men with a small AAA would lead to a 29% overall absolute risk reduction in 10-year predicted cardiovascular-events with seven cardiovascular-disease-free years of survival gained (6, 36).

It is not clear if using a different intervention development framework, such as the Behaviour Change Wheel (37) would result in a different outcome (37). However, both approaches require an initial identification of what behaviour(s) require changing, followed by identification of mechanisms /processes of behaviour change (including contextual /implementation factors) and appropriate behaviour change techniques that could be used to modify the identified change processes.

This is the first study to set out the theoretical framework and development process behind a cardiovascular risk reduction intervention in individuals with AAA or similar life-threatening vascular pathology. However, despite the significant detail presented, it is unlikely that a different team would exactly replicate this intervention, due to the interpretative nature of intervention development. The approach taken is robust and documentable however.

The following feasibility study will access the feasibility of using the developed intervention in routine clinical care. We will be able to fine-tune elements / components of the intervention or proceed to direct adoption into care, in discussion with relevant national stakeholders. All behavioural /psychological components of the intervention (our ‘targets for change’) are based on high-quality evidence (mostly of randomised nature), which has evaluated the clinical effectiveness of each parameter (e.g. weight loss, optimal blood pressure control, prescription of antithrombotic therapy and statins) for reducing cardiovascular risk.

In this work we present a detailed description of the design of an intervention to reduce cardiovascular risk in those with a AAA using intervention mapping (17). This intervention has been developed in collaboration with both stakeholders and patients, addressing the particular characteristics of both patients in AAA screening/surveillance across the NHS, as well as the actual clinical pathways in place. This intervention has the potential to save and improve thousands of lives in the future. Following a period of feasibility testing, the intervention will then be assessed in a trial, before wider adoption in the NHS.

## Data Availability

The data can be accessed via communication with the corresponding author

https://www.isrctn.com/ISRCTN93993995

## ACKNOWLEDGEMENTS

The research team are indebted to the imperative input of the patients and healthcare professionals in the design of this intervention.

## Licence statement

I, the Submitting Author has the right to grant and does grant on behalf of all authors of the Work (as defined in the below author licence), an exclusive licence and/or a non-exclusive licence for contributions from authors who are: i) UK Crown employees; ii) where BMJ has agreed a CC-BY licence shall apply, and/or iii) in accordance with the terms applicable for US Federal Government officers or employees acting as part of their official duties; on a worldwide, perpetual, irrevocable, royalty-free basis to BMJ Publishing Group Ltd (“BMJ”) its licensees and where the relevant Journal is co-owned by BMJ to the co-owners of the Journal, to publish the Work in BMJ Open and any other BMJ products and to exploit all rights, as set out in our licence.

The Submitting Author accepts and understands that any supply made under these terms is made by BMJ to the Submitting Author unless you are acting as an employee on behalf of your employer or a postgraduate student of an affiliated institution which is paying any applicable article publishing charge (“APC”) for Open Access articles. Where the Submitting Author wishes to make the Work available on an Open Access basis (and intends to pay the relevant APC), the terms of reuse of such Open Access shall be governed by a Creative Commons licence – details of these licences and which Creative Commons licence will apply to this Work are set out in our licence referred to above.

